# Same Inputs, Different EDSS: Measuring Specification Drift in Clinical Scoring Pipelines

**DOI:** 10.64898/2026.06.25.26356350

**Authors:** Sy Hwang, Sunil Thomas, Heather Williams, Amit Bar-Or, Vishakha Sharma, Frederik Buijs, Christopher Perrone, Danielle Mowery

## Abstract

Clinical informatics pipelines increasingly compute validated clinical endpoints from upstream NLP outputs. Even when the endpoint is defined by an established rubric, translating that rubric across representations—natural language instructions, program logic, and reference implementations—can introduce specification drift, where ostensibly equivalent calculators yield meaningfully different scores. We study this phenomenon for the Expanded Disability Status Scale (EDSS), a standard measure of disability in multiple sclerosis. Holding constant a shared set of functional-system (FS) subscores extracted by a large language model (LLM), we compare EDSS values computed across three representations of the same scoring rubric: prompt-executed natural language, LLM-generated code, and a canonical reference implementation. We characterize disagreement structure, distributional shifts, and clinically salient boundary flips, and we propose an audit workflow that treats endpoint computation as a first-class verification target in clinical NLP systems.

## Introduction

Clinical scoring rules are frequently treated as deterministic post-processing steps: once upstream variables are available, the endpoint follows mechanically. In practice, however, scoring rubrics are often encoded and deployed through a chain of translations—from published clinical descriptions to informal operational guidance, to prompts, to code—each of which may introduce ambiguity or interpretation. When such endpoints are used for longitudinal tracking, cohort selection, or outcome determination, even small systematic differences can cause downstream clinical or research consequences.

This paper studies specification drift in the computation of the Expanded Disability Status Scale (EDSS), a widely used and clinically significant disability measure in multiple sclerosis (MS). EDSS integrates impairment across functional systems and ambulation into a single ordinal score. Because EDSS is often used as a marker of progression and as a criterion for treatment decisions and clinical trial endpoints, robust and reproducible computation is important.

We evaluate three EDSS calculators that are intended to implement the same clinical rubric but differ in representation: a prompt-based natural-language calculator (NL-spec), an executable Python calculator (Exec-spec), and a canonical open-source comparator (Canonical). Crucially, all three calculators receive the same eight FS subscores predicted by an LLM from clinical notes. By isolating the EDSS conversion step, we aim to answer:

*How much endpoint variability is introduced purely by implementation representation, even when upstream inputs are identical?*

We make four contributions:

1. We formalize specification drift for clinical scoring as a measurable property of endpoint calculators.
2. We compare NL-spec, Exec-spec, and Canonical EDSS calculators under shared LLM-predicted FS inputs, quantifying disagreement patterns and distributional shifts.
3. We introduce clinically salient boundary flip analyses to interpret endpoint disagreement in practical terms.
4. We propose an audit workflow for endpoint computation that can be applied beyond EDSS to other clinical rubrics used downstream of NLP.

### Background and Related Work

#### EDSS as a computed clinical endpoint and precursor work

EDSS is a widely used ordinal measure of disability in MS, derived from examination-based FS subscores and ambulation, originally formalized by Kurtzke.^1^ Because EDSS is commonly used for longitudinal tracking of disability, cohort characterization, and research endpoints, reproducible computation is important, particularly near clinically salient boundary values where small score differences can change category membership or downstream operationalization.

Our study is a direct follow-on to our previous work demonstrating the feasibility of using LLMs to derive MS progression assessments from clinical notes, including FS subscores and EDSS.^2^ In that precursor work, we treated note-to-FS and FS-to-EDSS inference as an end-to-end clinical NLP problem and evaluated agreement for both intermediate subscores and final EDSS estimates. During subanalyses, however, we observed that EDSS values can be meaningfully sensitive to how the FS-to-EDSS conversion rules are instantiated, even when the upstream FS predictions are held fixed. This observation motivates the present study, which isolates endpoint computation and evaluates specification drift across the three calculator representations described below.

### Computable phenotypes, portability, and representation

EDSS conversion from FS subscores is a specific instance of a broader phenomenon of computable phenotypes and clinical scoring algorithms implemented over EHR data. The phenotyping literature has repeatedly emphasized that phenotype definitions must be shareable, interpretable, and executable across sites and data models, while supporting both human-readable and machine-executable forms.^3,4^ Pattern analyses of phenotype algorithms and their building blocks have likewise documented recurring design elements (structured queries, temporal logic, terminology constraints, and text/NLP components) that complicate portability.^5^

Multi-site experiences in the eMERGE network show that phenotypes with NLP/ML components can be portable, but require careful engineering choices, documentation, and accommodations for local variation.^6^ These findings motivate treating downstream scoring and aggregation logic as a first-class artifact. Even when upstream extraction is fixed, endpoint computation can drift as representations change.

### Differential and property-based testing for cross-implementation auditing

To systematically identify where implementations diverge, we draw from established software testing paradigms. Differential testing compares outputs across multiple implementations given identical inputs to uncover inconsistencies.^7^ Property-based testing generates many randomized inputs subject to constraints and checks whether specified properties hold.^8^ Modern implementations additionally support input shrinking to produce minimal counterexamples that are easier to interpret and regression-test.^9^ Conformance testing frameworks formalize the idea of checking whether an implementation meets specification requirements, often with structured coverage of the input space.**^?^**

In our context, these ideas translate naturally: we generate plausible FS vectors and use cross-implementation comparison to surface counterexamples across the three EDSS calculators. This provides a principled way to localize drift regions beyond what is observed in a finite clinical corpus.

### LLM-based execution and LLM-generated code

Recent work has demonstrated that large language models can synthesize executable code from natural language descriptions at useful levels of functional correctness.^10^ This capability enables new development patterns in which LLMs produce runnable implementations often through tool-augmented or agentic workflows, but it also raises concerns about correctness, stability, and verification. Our study leverages this setting as a natural experiment in whether prompt-executed and LLM-generated executable representations conform to an established canonical library.

## Methods

### Overall pipeline and study design

Clinical notes were first processed by an LLM to produce eight functional system (FS) subscores, including ambulation. These shared FS outputs were then passed, without modification, to the three EDSS calculators described below. The study was designed to isolate the EDSS conversion step: all three calculators consumed the same note-level FS vector, so observed differences reflect variation in endpoint computation rather than variation in upstream FS extraction.

### Inputs: LLM-predicted functional-system subscores

For each note, we used eight LLM-predicted FS subscores as fixed inputs: ambulation, visual, brainstem, pyramidal, cerebellar, sensory, bowel/bladder, and cerebral. These FS values were treated as fixed for all downstream analyses. We do not assume these FS subscores represent clinician-annotated ground truth. Rather, the aim of this study was to quantify the variability introduced by the EDSS conversion layer given identical upstream NLP outputs.

### EDSS calculators

**(1) NL-spec: prompt-based natural language specification** The NL-spec calculator provided a neurologist-authored stepwise description of the EDSS rubric, including pre-conversion rules and conditional scoring logic, to an LLM and requested a single EDSS output. NL-spec EDSS conversion was run through an OpenAI endpoint using openai-gpt-5; in this endpoint, temperature was not exposed as a configurable inference parameter. This representation operationalizes the rubric as natural language instructions rather than executable code. The full system prompt and inference configuration are provided in Appendix D.
**(2) Exec-spec: executable specification** The Exec-spec calculator was implemented as a deterministic Python realization of the same rubric. The Exec-spec calculator was developed using an LLM-assisted coding workflow but was executed deterministically after finalization. The agent was provided with the same neurologist-authored EDSS rubric used for the NL-spec prompt and asked to synthesize a Python function accepting the eight FS subscores as typed inputs and returning a single EDSS value. During development, the agent iteratively generated code, executed unit tests, inspected failures, and revised the implementation. Human review was used to verify that the final implementation encoded the intended pre-conversion rules, branch ordering, and output scale. After this development phase, the Python rules engine was frozen and applied deterministically to all FS vectors, with no LLM calls made during Exec-spec inference.
**(3) Canonical: open-source Kurtzke implementation** The Canonical calculator was a Python port of the published open-source adobrasinovic/edss implementation of Kurtzke EDSS logic (https://github.com/adobrasinovic/edss, src/index.js). We treated this implementation as a stable external reference for comparison, while recognizing that it also encodes specific interpretation choices.

### Comparison framework

We compared the three calculators using pairwise exact agreement, agreement within ±0.5 EDSS, and boundary-flip analyses at clinically relevant thresholds. Boundary flips were defined as cases in which two calculators placed the same note on opposite sides of a threshold. We also evaluated broader category reclassification using grouped EDSS ranges. EDSS values were compared on their native 0.5-step scale. Rows with negative FS subscores (n=8) were excluded only from analyses requiring valid non-negative FS vectors, such as witness grounding; pairwise agreement statistics were computed on non-missing calculator outputs.

### Algorithm divergence analysis

To characterize why deterministic implementations diverged, we performed exhaustive FS-space enumeration over ambulation scores 0–2 and all valid combinations of the remaining FS subscores. We then identified concrete FS vectors for which the Exec-spec and Canonical calculators produced different EDSS values. Minimal witness cases were used to localize disagreement to specific rule patterns, including high-FS edge cases, secondary-score tie-breaking, and interactions between ambulation-based and FS-based scoring logic.

### Agreement and drift metrics

Let *E*^(*a*)^ denote the EDSS computed for note *i* by calculator *a* ∈ {NL, Exec, Canon}.

#### Exact agreement

We compute pairwise exact match rates:

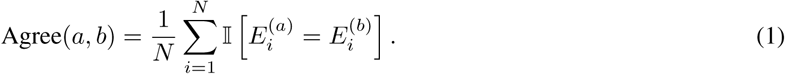

#### Tolerance agreement

Because EDSS is ordinal and reported in 0.5 increments, we also compute agreement within ±0.5:

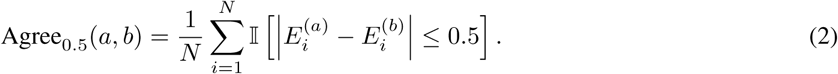

#### Distributional drift

We compare marginal distributions via the distance measure 1-Wasserstein distance.

#### Boundary-flip analyses

To interpret score differences operationally, we define boundary events *B*(*E*) such as:

- Threshold neighborhoods: indicator of being near key EDSS values (e.g., within 0.5 of 6.0).
- Category membership: rule-based bins reflecting disability strata (e.g., “≤ 5.5”, “6.0–6.5”, “≥ 7.0”).

A boundary flip occurs when two calculators place the same instance into different categories:

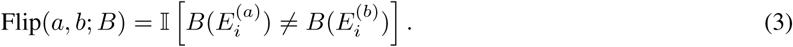

We report flip rates overall and stratified by EDSS range.

### Synthetic FS differential testing

To probe disagreement regions systematically, we generate plausible synthetic FS vectors under simple range constraints and compute EDSS across calculators. We then mine differential vectors where outputs disagree and cluster them by ambulation strata and non-ambulatory FS severity patterns. This produces minimal counterexamples that help localize which logical branches differ across implementations.

## Experiments

### Dataset

We evaluate on a cohort of clinical notes from patients in the Penn Neuroimmunology Registry with a diagnosis of MS by a neurologist according to McDonald criteria.^11^ The dataset totaled 57,604 progress notes across 5,231 patients. We analyze EDSS at the note level and optionally summarize at the patient level for longitudinal plausibility checks.

### Experimental protocol

1. Extract FS subscores from each note using a fixed LLM configuration.
2. Compute EDSS using NL-spec, Exec-spec, and Canonical calculators on the identical FS vectors.
3. Quantify pairwise agreement, tolerance agreement, and marginal distribution differences.
4. Compute boundary-flip rates, overall and stratified by EDSS range.

## Results

### Agreement and distributional drift

Table 1 reports exact and tolerance agreement between calculators, and Figure 1 compares EDSS distributions across calculators. The strongest agreement was between Exec-spec and Canonical, which matched exactly for 98.1% of notes and were within ±0.5 EDSS for 99.8%. In contrast, comparisons involving NL-spec showed lower exact agreement: 84.2% against Exec-spec and 82.8% against Canonical, although roughly 90% of notes remained within one half-step.

**Figure 1:**
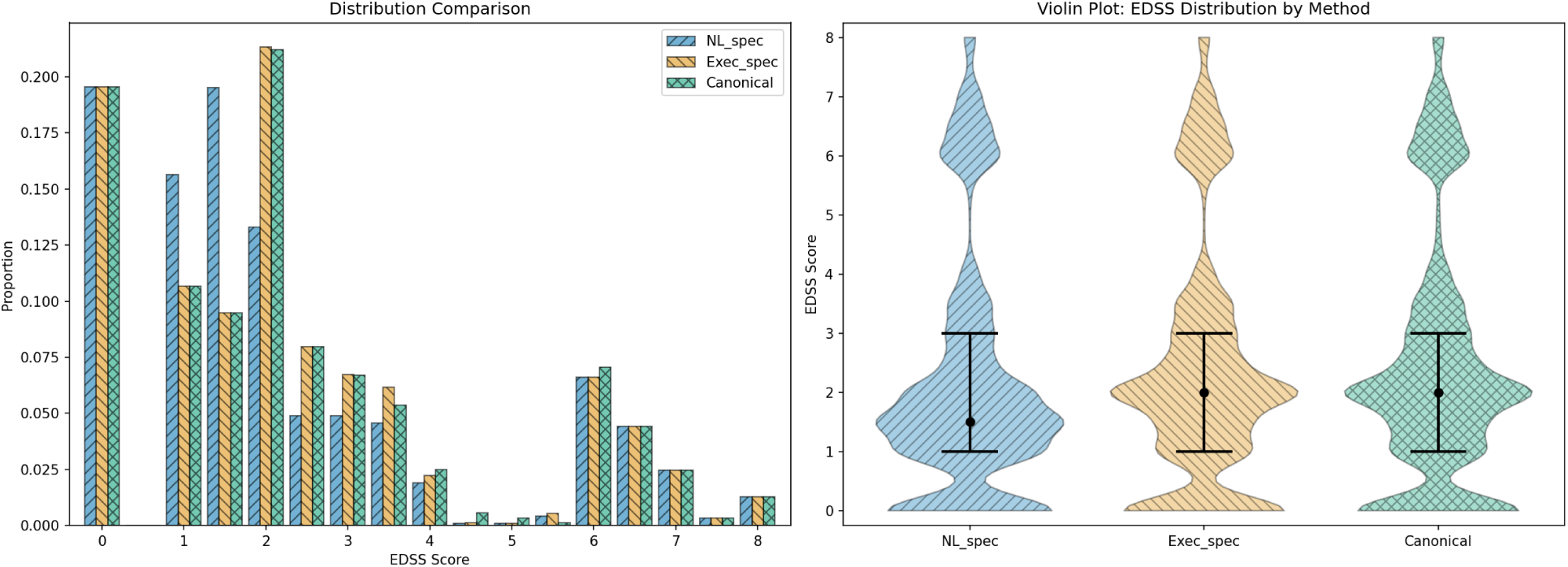
Distribution comparison of EDSS across calculators. Pronounced differences in the EDSS 1–4 range indicate representation-driven drift in endpoint computation despite identical FS inputs.

**Table 1:** Pairwise agreement between EDSS calculators on complete-case observations (*n* = 57,604). Exact agreement requires identical EDSS; tolerance agreement counts outputs within ±0.5.

| Pair | Exact agreement (%) | $\pm 0.5$ agreement (%) | 1-Wasserstein |
| --- | --- | --- | --- |
| NL-spec vs Exec-spec | 84.2 | 90.2 | 0.170 |
| NL-spec vs Canonical | 82.8 | 90.1 | 0.183 |
| Exec-spec vs Canonical | 98.1 | 99.8 | 0.014 |

The distributional results followed the same pattern. Exec-spec and Canonical had minimal marginal drift (1-Wasserstein distance 0.014), whereas NL-spec differed more substantially from both deterministic comparators (0.170–0.183). Visual inspection of the score distributions indicated that these differences were concentrated primarily in the EDSS 1–4 range. Thus, specification drift was not evenly distributed across the scale; it was concentrated in the mild-to-moderate region where multiple FS-based rule branches can plausibly apply. Appendix A reports the corresponding calculator-vs-annotator robustness checks on the clinician-annotated subset, and Appendix E reports repeat-call NL-spec reliability analyses.

### Synthetic divergence witnesses

Synthetic FS differential testing localized deterministic disagreement between Exec-spec and Canonical to a small set of rule patterns. Exhaustive enumeration over low-ambulation FS vectors produced seven minimal divergence patterns, dominated by cases involving high non-ambulatory FS scores (≥ 5), second-largest FS tie-breaking when the maximum FS score was 3 or 4, and precedence between ambulation-based and FS-based scoring (Table 2). Although some enumerated witnesses were clinically implausible, five of seven divergence patterns were represented in the observed cohort; 2.39% of notes shared a witness rule pattern and 1.45% exactly matched a synthetic witness vector. These findings indicate that synthetic differential testing did not merely identify theoretical edge cases, but exposed rule regions reachable in real clinical data. Appendix C reports the full grounding analysis.

**Table 2:** Algorithm divergence between the Exec-spec and Canonical EDSS calculators. Each row represents a minimal FS subscore vector that produces different EDSS scores. Divergence patterns for ambulation 0 and 1 are identical; rows marked “0–1” apply to both.

| FS Subscores |  |  |  |  |  |  |  |  |  |  |  |  |  |  |
| --- | --- | --- | --- | --- | --- | --- | --- | --- | --- | --- | --- | --- | --- | --- |
| Amb. | Vis | Bs | Pyr | Cer | Sen | BB | Cb | Pattern |  | Exec | Canon | Diff | Exec-spec explanation | Canonical explanation |
| 0–1 | 0 | 0 | 0 | 0 | 0 | 0 | 5 | 1×FS=5 |  | 2.0 | 5.0 | +3.0 | Does not recognize that one system has a very severe score (5); treats this as a low-disability case by default | Detects that the highest system score is 5 or above and immediately assigns 5.0 |
| 0–1 | 0 | 0 | 0 | 0 | 0 | 3 | 5 | 1×FS=5, 1×FS=3 | | 3.0 | 5.0 | +2.0 | Ignores the system scored 5; only sees one system at 3 with none at 2, yielding 3.0 | Detects a system score $\geq 5$ and immediately assigns 5.0, regardless of other scores |
| 0–1 | 0 | 0 | 0 | 0 | 2 | 3 | 3 | 2×FS=3, 1×FS=2 |  | 3.5 | 4.0 | +0.5 | Sees two systems at 3 and assigns 3.5; does not consider what the next-highest score is | Also sees two systems at 3, but additionally checks the next-highest score; because it is 2, escalates to 4.0 |
| 0–1 | 0 | 0 | 0 | 0 | 2 | 3 | 5 | 1×FS=5, 1×FS=3, 1×FS=2 | | 3.5 | 5.0 | +1.5 | Ignores the system scored 5; sees one system at 3 and one at 2, yielding 3.5 | Detects a system score $\geq 5$ and immediately assigns 5.0 |
| 0–1 | 0 | 0 | 0 | 0 | 0 | 2 | 4 | 1×FS=4, 1×FS=2 |  | 4.0 | 4.5 | +0.5 | Sees one system at 4 and assigns 4.0; does not consider what the next-highest score is | Also sees one system at 4, but additionally checks the next-highest score; because it is 2, escalates to 4.5 |
| 0–1 | 0 | 0 | 0 | 0 | 0 | 5 | 4 | 1×FS=5, 1×FS=4 |  | 4.0 | 5.0 | +1.0 | Sees at least one system at 4 and assigns 4.0; does not distinguish having one vs. two systems at that level | Counts two systems at 4 after bowel/bladder conversion from 5 to 4 and escalates to 5.0 |
| 2 | 0 | 0 | 0 | 0 | 0 | 0 | 5 | 1×FS=5 | | 4.5 | 5.0 | +0.5 | With ambulation = 2, directly assigns 4.5 based on walking distance alone, ignoring all system scores | Checks system scores first; detects a score $\geq 5$ and assigns 5.0, overriding the ambulation-based rule |

### Boundary flips and practical impact

We next interpret drift through category flips that would matter operationally. Table 3 and Figure 2 report flip rates for clinically meaningful EDSS decision boundaries rather than only raw score disagreement. These boundaries correspond to transitions between broad disability states with distinct implications for interpretation and downstream use. In particular, crossing from EDSS 3.0–5.5 to EDSS 6.0–7.5 is especially important because it marks loss of independent ambulation and entry into assistance- or wheelchair-dependent disability states, while movement into EDSS 8.0+ reflects severe disability with major dependence in daily functioning.

**Figure 2:**
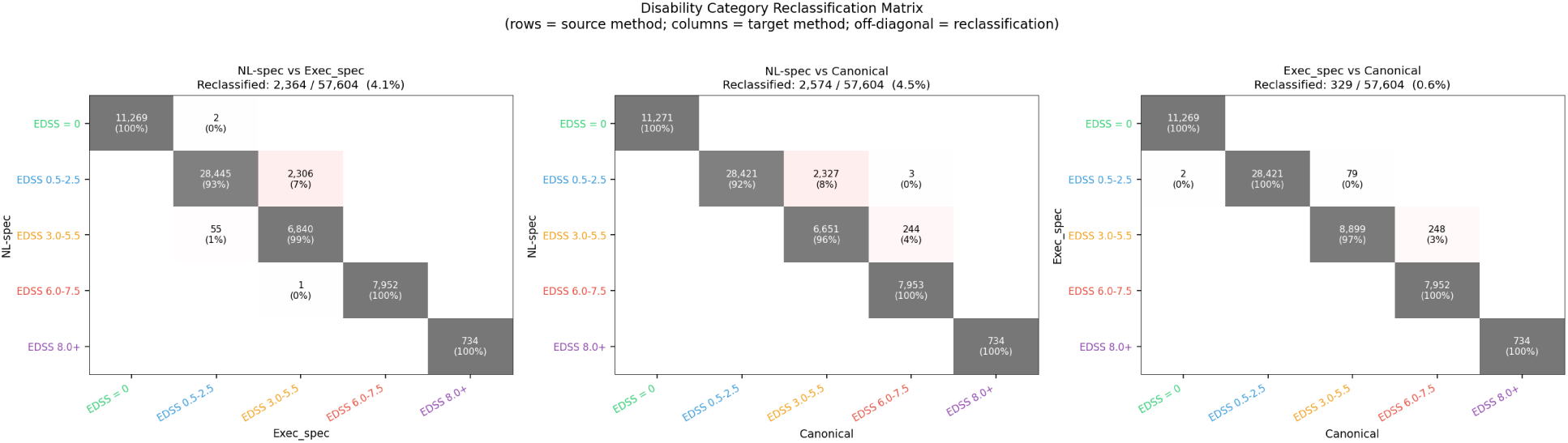
Boundary-flip reclassification matrices for pairwise EDSS comparisons: NL-spec vs Exec-spec (left), NL-spec vs Canonical (middle), and Exec-spec vs Canonical (right). Each cell shows the number of complete-case observations reclassified across clinically meaningful disability strata (EDSS 0–3.5, 4.0–5.5, 6.0–7.5, and 8.0+). Darker cells indicate higher concentrations of discordant assignments.

**Table 3:**
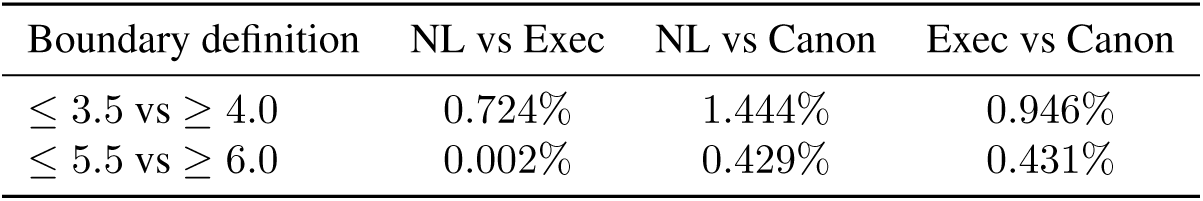
Boundary-flip rates: proportion of notes whose category membership changes between calculators.

| Boundary definition | NL vs Exec | NL vs Canon | Exec vs Canon |
| --- | --- | --- | --- |
| $\leq 3.5$ vs $\geq 4.0$ | 0.724% | 1.444% | 0.946% |
| $\leq 5.5$ vs $\geq 6.0$ | 0.002% | 0.429% | 0.431% |

Viewed this way, a “boundary flip” is a potential reclassification into a different clinically interpreted severity stratum. This framing is more relevant for practice than exact-score agreement alone, because many downstream applications, including severity stratification, cohort definition, and progression analyses, depend on whether a patient falls on one side or the other of such thresholds. Low flip rates at higher-severity boundaries would suggest that disagreements are concentrated in milder disease ranges and are less likely to alter major disability-state assignments, whereas elevated flip rates near a given threshold would indicate greater risk of clinically meaningful reclassification at that boundary. Threshold-specific flip counts across candidate EDSS cut points are provided in Appendix B.

## Discussion

Our results show that EDSS values computed from identical FS inputs can differ meaningfully across calculator representations. This indicates that endpoint computation itself can be a nontrivial source of variability in clinical NLP pipelines, independent of upstream extraction error.

### Why drift arises

We observed several plausible mechanisms for representation-driven drift across the three EDSS calculators:

- Ambiguity in natural language rules: when rules are expressed in prose, overlapping conditions can remain under-specified. For example, statements such as “one FS = 3” and “multiple FS = 2” may both apply to the same input pattern, but the intended precedence is not always explicit. An LLM executing the NL-spec may therefore resolve the same rule set differently from a hand-coded implementation.
- Implicit tie-breaking and branch ordering: deterministic implementations must choose an explicit evaluation order, even when the underlying clinical description does not specify one. In our comparison, divergence often occurred in cases where multiple clauses were simultaneously satisfiable, such as when the maximum FS score was 3 or 4 and secondary FS scores were also elevated. The canonical implementation used additional context whereas the executable specification often relied on simpler count-based rules, producing systematic 0.5-point differences.
- Pre-conversion logic and threshold handling: EDSS calculation depends not only on raw FS subscores but also on score normalization steps, such as visual and bowel/bladder conversions. Small differences in how these conversions are encoded can shift downstream branch selection. More importantly, edge-case thresholds can be omitted or treated differently across representations. In our exhaustive comparison, several of the largest disagreements arose when one implementation explicitly handled high FS values while another fell through to lower-score count-based logic.
- Ambulation-versus-FS precedence: another recurrent source of drift was whether ambulation should dominate the final EDSS assignment or whether sufficiently severe non-ambulatory FS scores should override the ambulation-based score. For example, the executable specification mapped some ambulation scores directly to EDSS values, whereas the canonical implementation first checked for severe FS abnormalities and escalated the score accordingly. This produced clinically meaningful disagreements even when the absolute difference was only 0.5–1.0 points.
- LLM behavior as an interpreter: NL-spec execution by an LLM is not equivalent to symbolic rule execution. Even with a fixed prompt, the model may compress multiple clauses into a heuristic mapping, smooth over rare edge cases, or privilege common score patterns seen during pretraining over strict rule precedence. As a result, the NL-spec can behave more like a learned approximation of the rules than a literal interpreter of them.
- Representation-specific failure modes: each representation introduces its own error profile. Natural language specifications are vulnerable to ambiguity, executable specifications to implementation omissions, and canonical external libraries to hidden assumptions inherited from prior codebases. The observed drift is therefore not attributable to a single bug or model error, but to the fact that the same clinical logic is being instantiated in different representational forms.

The minimal deterministic witness cases in Table 2 provide concrete FS vectors illustrating several of these mechanisms.

### Implications for clinical informatics and governance

Even in the absence of clinician-annotated FS subscores, our findings highlight a governance-relevant principle for clinical informatics: computable endpoints should be managed as first-class artifacts, not treated as static labels or informal prompt outputs. In practice, once a derived endpoint such as EDSS is used for cohort definition, severity stratification, progression analyses, treatment comparisons, or downstream modeling, the implementation itself becomes part of the study instrument. Small changes in how the endpoint is represented can alter category assignments, measured agreement, and analytic conclusions.

For EDSS-like rubrics, this argues for a layered governance strategy. First, operational use should be anchored to a canonical executable implementation that is versioned and reviewable, rather than to prose alone. Natural-language specifications are valuable for communicating intent, but they do not by themselves guarantee reproducible execution. Second, executable implementations should be accompanied by regression tests that cover both common cases and edge cases, including minimal witness cases that expose known divergence modes. In our study, exhaustive FS-space enumeration made it possible to identify a small number of concrete rule patterns responsible for disagreement; these witness cases can be retained as permanent test fixtures whenever the endpoint logic is updated. Third, teams should maintain explicit documentation of interpretation choices, including branch ordering, tie-breaking, score conversion rules, and handling of rare or underspecified boundary conditions. These choices are often clinically consequential precisely because they appear minor at the code level.

More broadly, our results suggest that endpoint governance for LLM-enabled pipelines should mirror governance already expected for clinical decision rules, phenotypes, and computable quality measures. This includes version control, change logs, unit/regression testing, reference outputs, and periodic revalidation when prompts, model endpoints, or external libraries change. Without such controls, representational drift can be mistaken for patient-level signal, especially when an endpoint is re-used across studies or over time.

LLM-based calculators remain useful, particularly during rapid prototyping, rubric translation, or exploratory analyses where the goal is to test feasibility before investing in a full executable specification. However, they should be treated as a candidate implementation rather than as the authoritative endpoint definition. Before operational deployment or inferential use, they should be benchmarked against a canonical executable reference and monitored for drift under prompt, model, or vendor changes. In this sense, the central governance lesson is not that LLMs should be excluded from endpoint generation, but that their outputs should enter the same verification, versioning, and auditing framework expected of any other clinical computation.

### Generalization beyond EDSS

The proposed audit framework applies broadly to computed clinical endpoints and phenotyping rules such as severity indices and risk scores, when those endpoints are derived from NLP-extracted variables. As LLMs increasingly participate in both extraction and downstream computation, evaluation should include not only extraction accuracy but also endpoint computation fidelity.

### Limitations

This study has several limitations. First, we evaluated specification drift using LLM-predicted FS subscores rather than clinician-annotated FS ground truth. Accordingly, our analyses compare the behavior of alternative EDSS calculators under a shared input representation, but do not establish that any one calculator is uniformly more clinically correct than the others. Our primary claim is therefore about representation- and implementation-dependent variation in endpoint computation, not about absolute clinical validity of a particular EDSS implementation.

Second, the NL-spec calculator is implemented through an LLM and thus does not inherit the deterministic guarantees of a conventional rules engine. Even when prompts and model endpoints are held constant, repeatability is an empirical property rather than a formal one, and may change with model updates, serving infrastructure, or prompt revisions. Although our study treats the NL-spec as one operational realization of the natural language rules, it should be understood as a probabilistic interpreter rather than a symbolic executor. The repeat-call analyses in Appendix E estimate how much observed NL-spec disagreement is attributable to stochastic execution versus systematic representation drift.

Third, the canonical open-source implementation was treated as a stable reference for comparison, but it is not a gold standard in the sense of being interpretation-free. Like any executable implementation of a clinical rubric, it embodies specific assumptions about score conversion, precedence, and tie-breaking. Some disagreements identified in our analyses therefore reflect differences in encoded interpretation rather than a simple distinction between “correct” and “incorrect” implementations. In this sense, the canonical comparator is best viewed as a well-defined external reference, not as an unquestionable source of truth.

Fourth, our analyses focus on EDSS, a rubric with ordinal scoring, branch-based logic, and clinically meaningful thresholds. Other clinical scoring systems may exhibit different drift profiles depending on their structure. Rubrics with fewer branch points, no pre-conversion rules, or weaker threshold effects may show smaller representation-dependent differences, whereas more complex composite scores may show larger or less interpretable divergence. The general lesson about endpoint governance likely transfers, but the magnitude and clinical significance of drift will need to be established separately for other instruments.

Finally, our synthetic divergence analyses intentionally explored the full rule space, including combinations that may be rare or clinically implausible in practice. This was useful for identifying mechanistic sources of disagreement, but not all witness cases should be interpreted as equally likely in real-world cohorts. For this reason, we paired exhaustive enumeration with cohort-based agreement, boundary-flip analyses, and the witness-grounding analysis in Appendix C to distinguish theoretical divergence from practically observed divergence.

### Conclusion

We present a focused study of specification drift in EDSS computation across three representations of the same scoring rubric. Using shared FS inputs, we show that calculator disagreement is not random noise but a structured phenomenon concentrated in particular rule regions and, in some cases, associated with clinically relevant boundary flips. Even when absolute score differences are modest, those differences can alter category assignment and therefore affect downstream cohorting, progression analyses, and interpretation of disease severity.

More broadly, our findings argue that endpoint computation should be treated as a verification target in clinical NLP systems. For composite clinical scores, it is not sufficient to report model performance only at the extraction layer; one must also assess how extracted values are transformed into derived endpoints. We therefore recommend an audit workflow that combines pairwise agreement analysis, boundary-flip analysis, and exhaustive witness-case testing to identify both the impact and the mechanism of specification drift. In operational settings, this supports a governance model in which endpoint calculators are versioned, regression-tested, and explicitly documented with respect to tie-breaking and edge-case handling.

The larger implication is that representational form matters. A prose rubric, an executable rules engine, and an external reference library may all appear to implement “the same” clinical endpoint while still producing materially different outputs. Making those differences visible, auditable, and testable is an important step toward more reliable and governable computed endpoints in clinical informatics.

## Data Availability

The clinical note data analyzed in this study are not publicly available due to patient privacy and institutional data-use restrictions. Additional derived data may be available from the corresponding author upon reasonable request and subject to institutional approvals.

## Acknowledgments

This project was partially funded by a grant from F. Hoffmann-La Roche, Basel, Switzerland.

## A Calculator-vs-annotator robustness

We compared NL-spec, Exec-spec, and Canonical calculators against the pooled annotator EDSS on exact accuracy, lenient agreement, quadratic weighted kappa (QWK), mean absolute error (MAE), signed bias, and within-tolerance agreement. This analysis evaluates whether the choice of EDSS calculator materially changes performance conclusions on the clinician-annotated subset.

**Table 4:** Three-way EDSS-derivation comparison against pooled annotator EDSS.

| Calculator | Exact acc. | Lenient acc. | QWK | MAE | Mean bias | Within 0.5 | Within 1.0 |
| --- | --- | --- | --- | --- | --- | --- | --- |
| NL-spec | 0.76 | 0.98 | 0.978 | 0.18 | -0.14 | 0.94 | 0.98 |
| Exec-spec | 0.80 | 0.98 | 0.980 | 0.15 | -0.11 | 0.96 | 0.98 |
| Canonical | 0.82 | 0.98 | 0.981 | 0.14 | -0.06 | 0.96 | 0.98 |

**Table 5:** Signed bias by EDSS calculator.

| Calculator | Mean signed error | 95% CI | % over | % under | % exact |
| --- | --- | --- | --- | --- | --- |
| NL-spec | -0.14 | (-0.26, -0.02) | 4% | 20% | 76% |
| Exec-spec | -0.11 | (-0.23, 0.01) | 4% | 16% | 80% |
| Canonical | -0.06 | (-0.18, 0.06) | 8% | 10% | 82% |

**Table 6:**
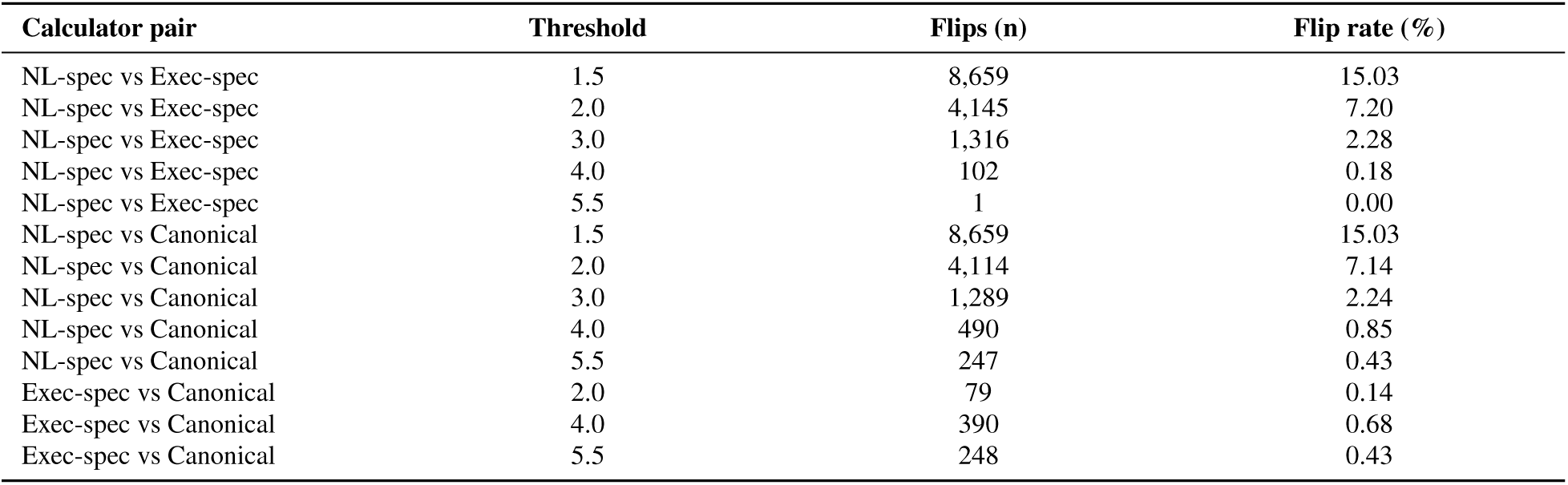
Boundary flips by threshold across the full cohort.

All three calculators achieved essentially identical lenient agreement (0.98) and QWK (∼0.98). The Canonical calculator had the lowest absolute bias (-0.06 EDSS) and the highest exact accuracy, supporting its use as the primary reference implementation in the drift analyses.

### B Boundary-flip analysis across candidate thresholds

For each pair of calculators, we counted the number and rate of notes whose derived EDSS fell on opposite sides of each candidate threshold from 1.5 through 6.5. This threshold-level view complements the broader disability-stratum reclassification matrix in the main text.

At clinically pivotal thresholds, including EDSS 4.0 and 5.5–6.0, reclassification between calculators was rare (≤0.85%). Exec-spec and Canonical agreed almost perfectly except in the EDSS 4.0–5.5 moderate range.

### C Grounding synthetic witness vectors in the observed cohort

Exhaustive synthetic witness testing enumerates every FS vector consistent with the EDSS coding manual and is therefore agnostic to clinical plausibility. To assess whether synthetic witnesses included FS combinations not seen clinically, we anchored every synthetic witness vector to the 57,604-note cohort (*n* = 57,596 with complete, non-negative FS values).

For every synthetic witness 8-tuple (amb*, V, Bs, Py, Ce, Se, BB, Cb*), we computed its exact observed frequency, its nearest-neighbor *L*_1_ distance to any observed FS vector using a *k*-d tree, and its rule-pattern frequency in the cohort. Witnesses were then grouped by rule pattern.

Of the seven distinct divergence patterns identified by synthetic enumeration, five were realized in the observed cohort with non-zero pattern frequency (Table 7). In aggregate, 2.39% of cohort notes shared a rule pattern with at least one synthetic witness, and 1.45% matched a synthetic witness FS vector exactly. The two absent patterns were clinically implausible combinations, such as ambulation already requiring a cane co-occurring with a non-ambulatory FS at level 5+, and together accounted for 26.9% of the synthetic witness pool.

**Table 7:**
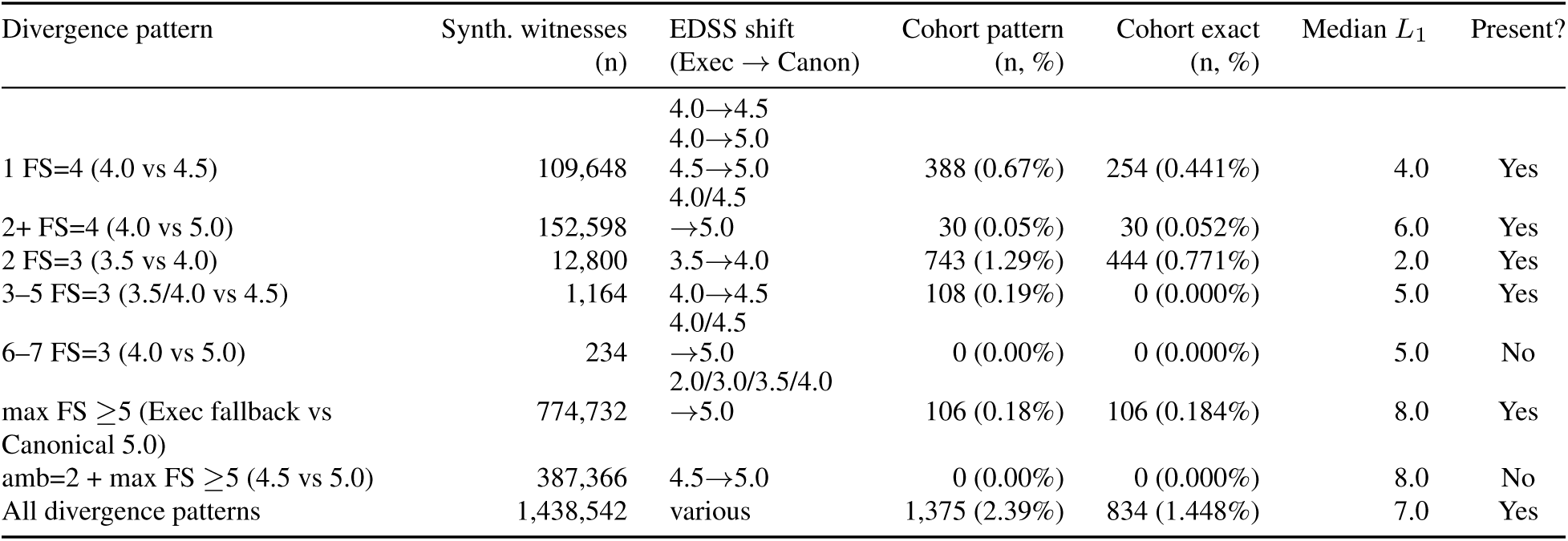
Per-pattern grounding of synthetic witness vectors against the observed cohort (*n* = 57,596). *Cohort pattern* = notes whose FS rule pattern matches the divergence pattern; *Cohort exact* = notes whose raw FS 8-tuple exactly matches a synthetic witness; *Median L*_1_ = nearest-neighbor Manhattan distance from synthetic witness to observed FS vector.

The most common drift patterns, including single FS=4 and multi-system FS=2/FS=3 profiles, were also the most prevalent in the clinical cohort, indicating that synthetic differential testing exercised clinically realistic regions of the rule space. Even for witnesses whose exact FS vector was unobserved, the median *L*_1_ distance to an observed vector remained small enough to show that the exposed drift cells are reachable from real clinical profiles by modest perturbations rather than being artifacts of exhaustive enumeration.

**Figure 3:**
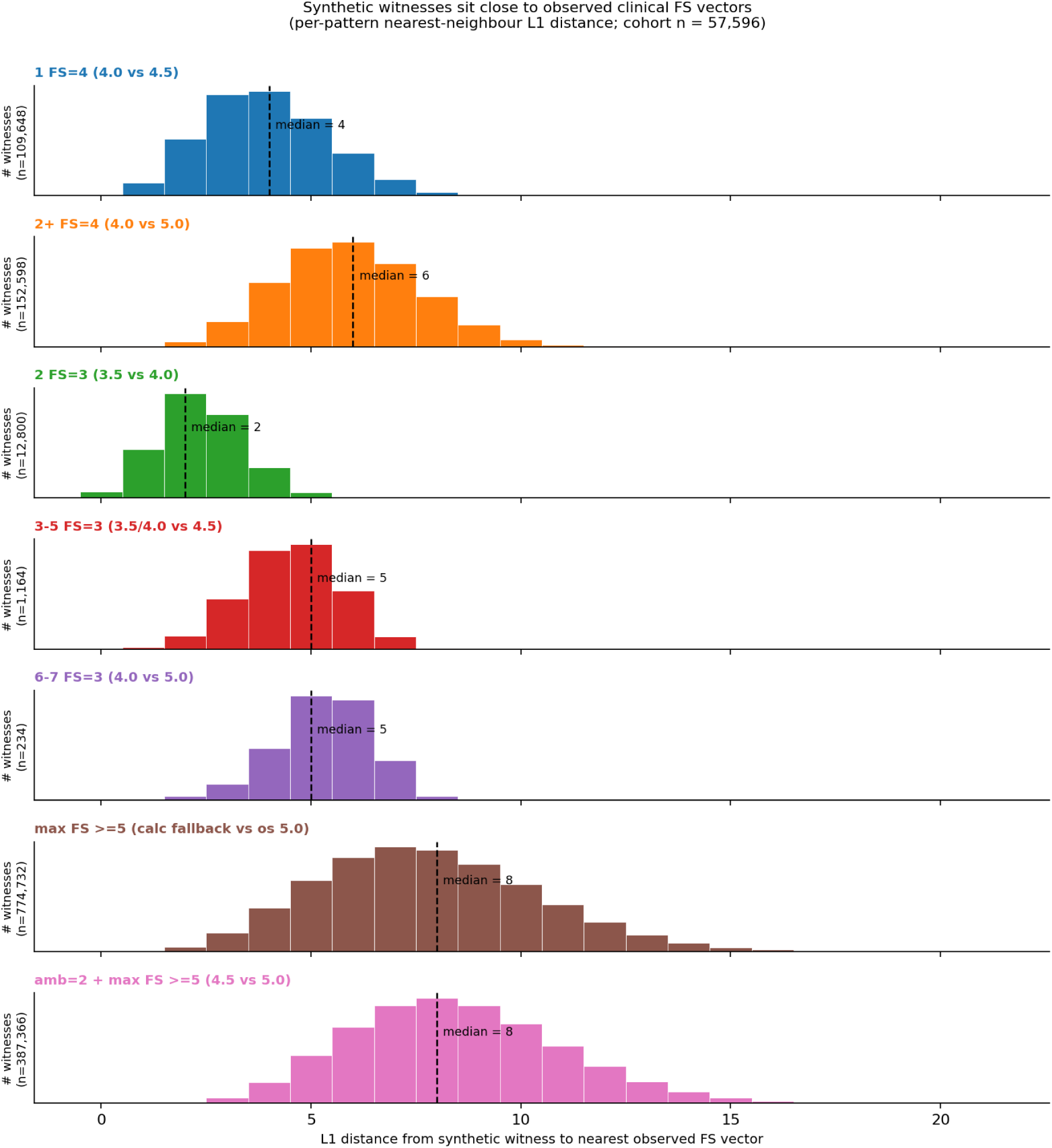
Per-pattern distribution of *L*_1_ nearest-neighbor distance from each synthetic witness 8-tuple to the closest observed FS vector in the cohort. Dashed lines mark per-pattern median distance.

### D NL-spec prompt and inference configuration

The NL-spec calculator was implemented as a prompt-executed natural-language specification. At inference time, the model received only the eight functional system (FS) subscores listed in the input prompt below; it did not receive the original clinical note text, clinician annotations, patient metadata, or outputs from the other EDSS calculators. The prompt did not include few-shot examples, and the model was instructed to return only the final EDSS value as a numeric score with one decimal place.

**Table 8:** Inference configuration for the NL-spec calculator.

| Setting | Value |
| --- | --- |
| Model | openai-gpt-5 |
| Reasoning effort | medium |
| Maximum tokens | 10,000 |
| Input context | Eight FS subscores only |
| Clinical note text provided | No |
| Patient metadata provided | No |
| Few-shot examples | No |
| Chain-of-thought requested | No |

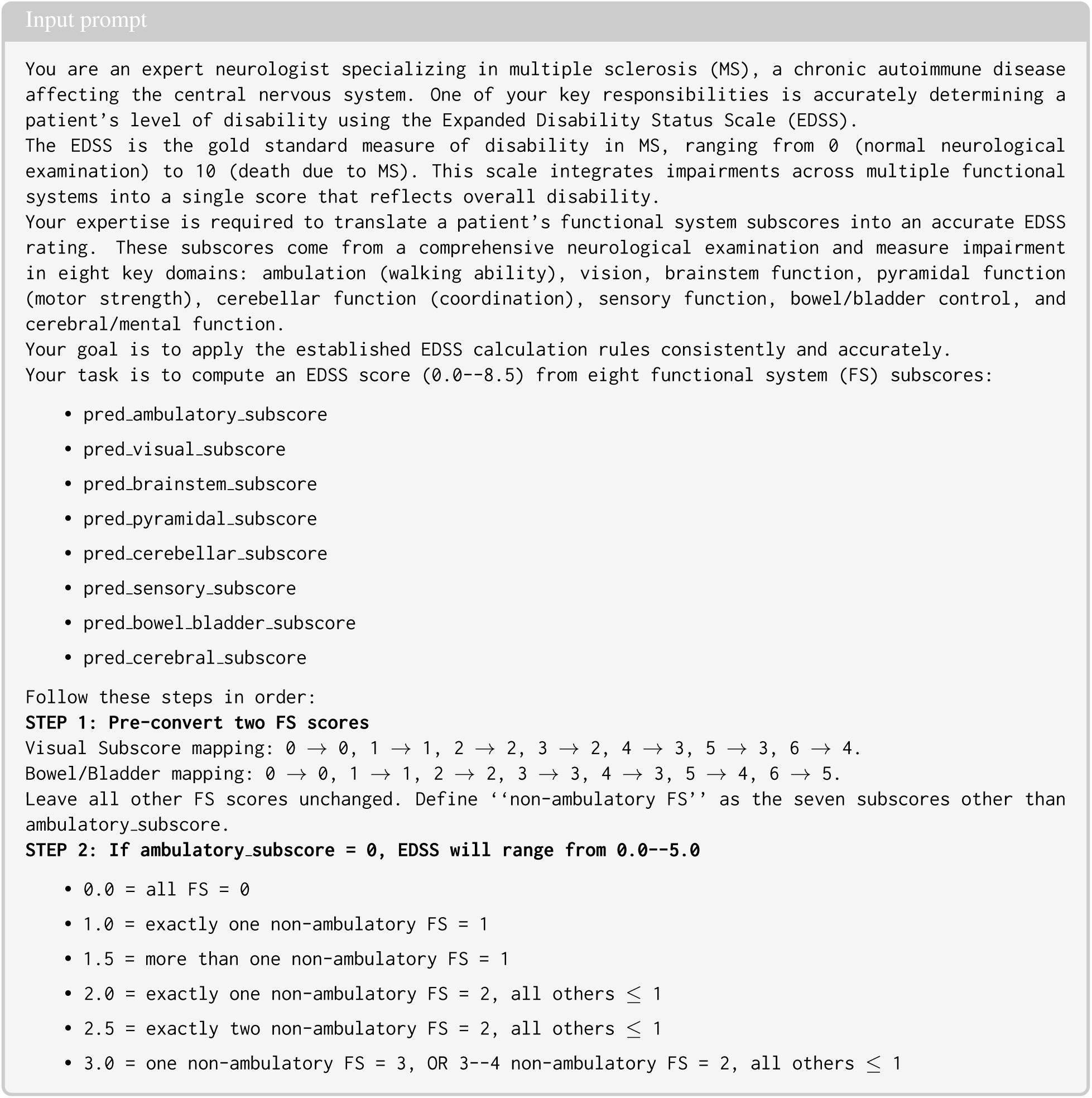
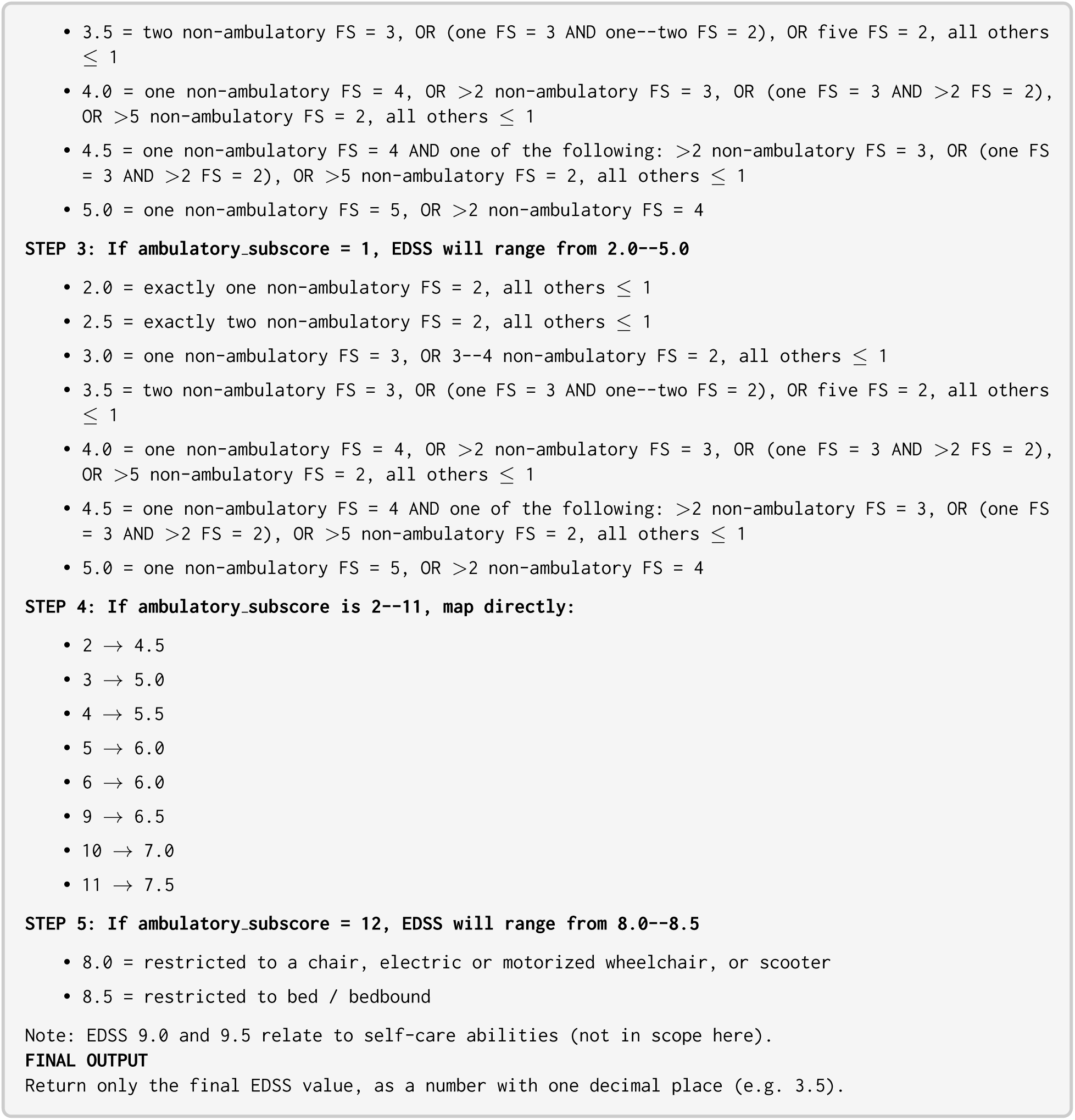

### E LLM reproducibility and test–retest reliability

Because LLMs are non-deterministic, some fraction of method disagreement could reflect call-to-call execution variability rather than systematic differences in how the natural-language specification encodes the EDSS algorithm. We evaluated this directly by scoring each note multiple times and comparing within-note variability against the between-method gap.

Each of the 57,604 notes was independently scored five times by GPT-5 (reasoning effort = medium). For each note, we computed a test–retest range [min, max] over the five replicates and a drift measure |median(replicates) − Exec-spec|, both in EDSS units. A note exhibited execution noise when the Exec-spec value fell inside the test–retest range and representation drift otherwise.

**Table 9:** Test–retest reliability and four-category partition across five GPT-5 replicates per note (*n* = 57,604).

| Reliability metric | Value | Partition category | n | % |
| --- | --- | --- | --- | --- |
| ICC(2,1), absolute agreement | 0.996 | Stable agreement | 38,861 | 67.5 |
| Mean pairwise exact agreement | 91.4% | Noise around Exec-spec | 9,637 | 16.7 |
| Mean pairwise MAE | 0.05 EDSS | Disagreement within range | 483 | 0.8 |
|  |  | Disagreement beyond range | 8,623 | 15.0 |

Among notes whose median NL-spec value differed from Exec-spec (*n* = 9,106; 15.8%), 94.7% remained disagreements across all five replicates with Exec-spec outside the test–retest range. Only 5.3% had a test–retest range containing the Exec-spec value. Among notes where a single call disagreed with Exec-spec, only 4.1% had at least one replicate land on the Exec-spec value, so re-querying would resolve fewer than one in twenty disagreements.

**Figure 4:**
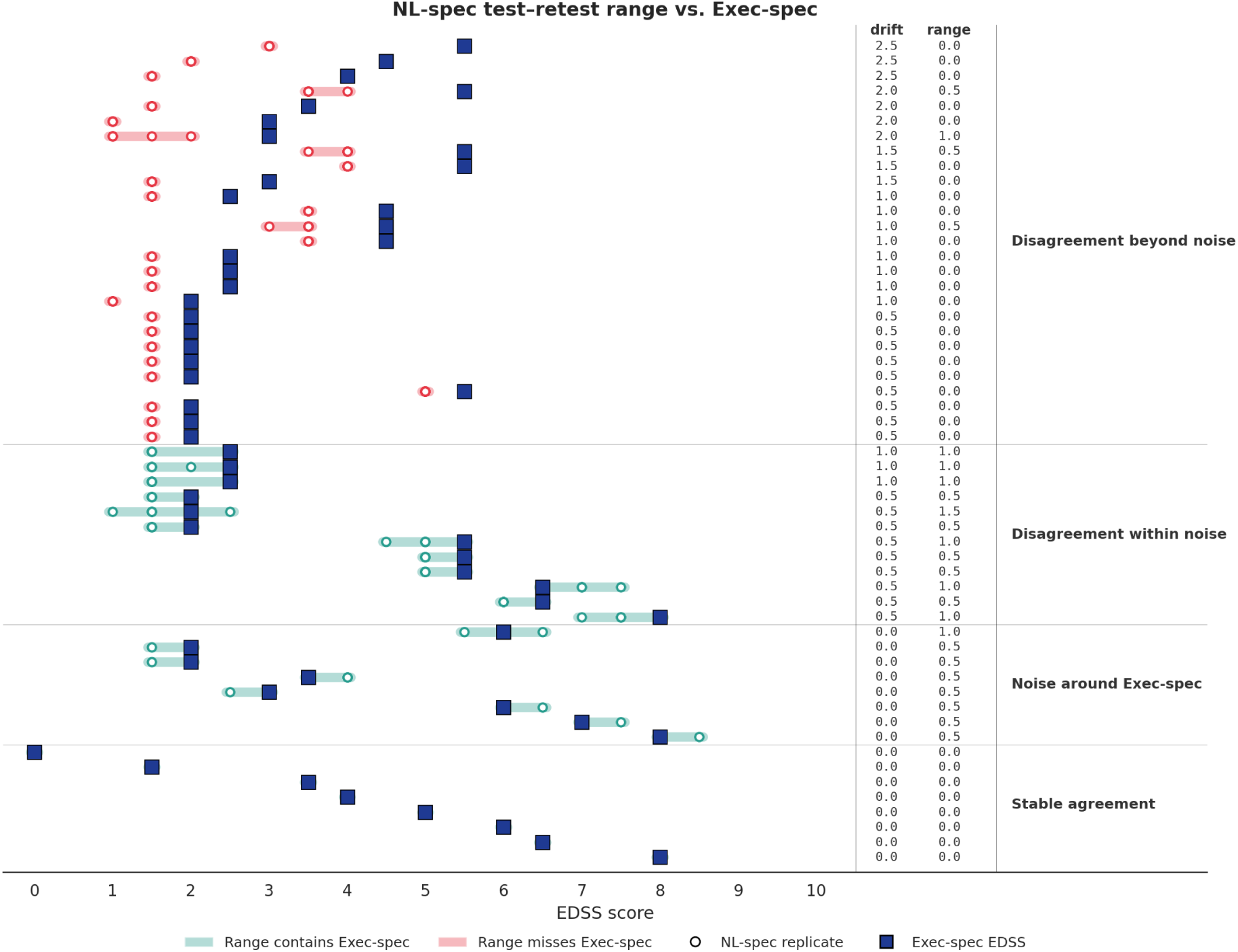
Per-note test–retest range vs. Exec-spec for a stratified sample of 60 notes. Horizontal bars span [min, max] across the five NL-spec replicates; open circles mark individual replicate values; filled squares mark deterministic Exec-spec scores.

**Figure 5:**
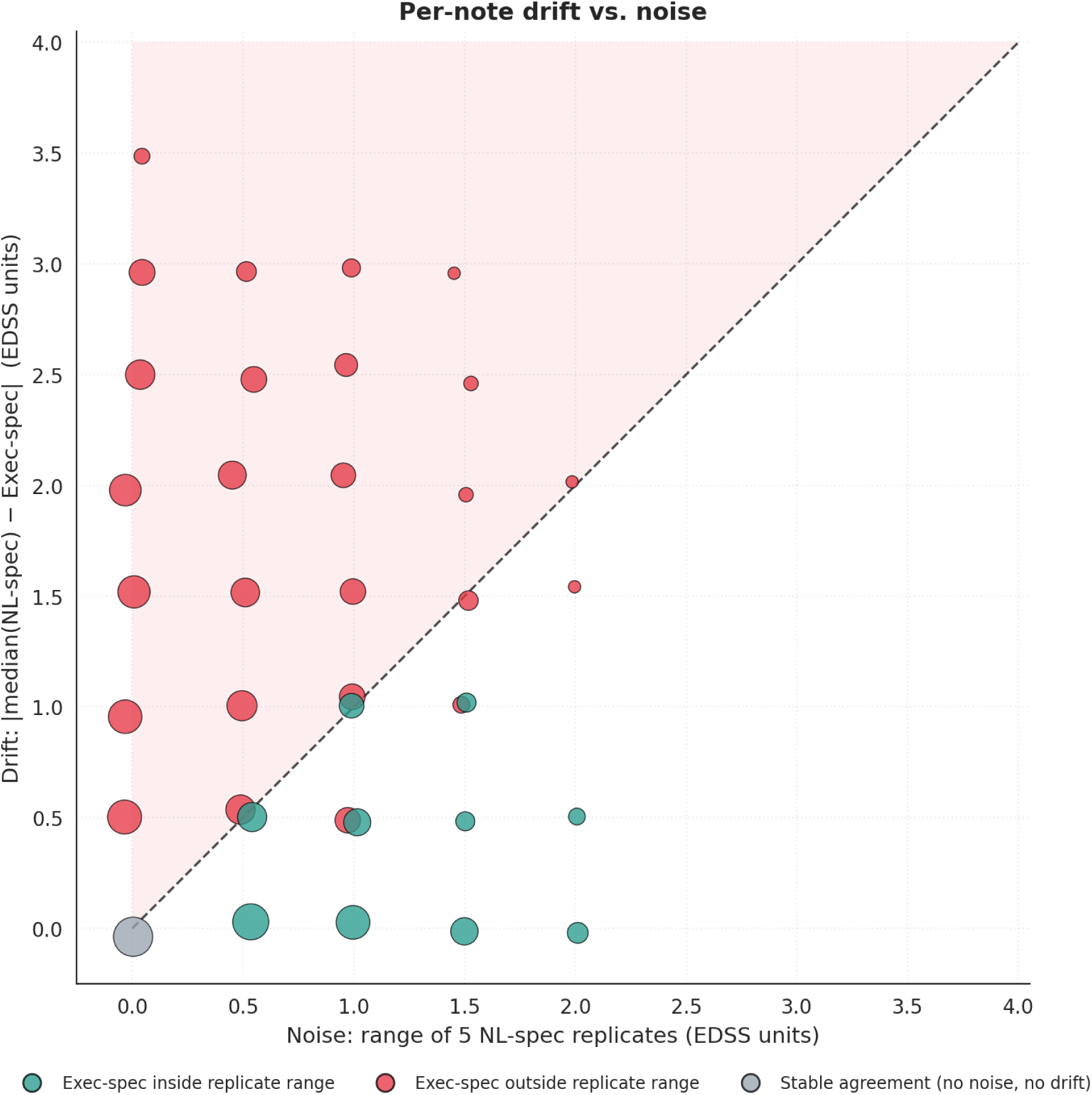
Per-note drift vs. noise across the full cohort. Points on or below the diagonal could in principle be reconciled by re-querying the LLM, whereas points above the diagonal cannot.

### F Within-prompt stochastic decomposition

To isolate within-prompt sampling noise from systematic disagreement, we repeated the NL-spec calculation on 120 FS vectors stratified into background, boundary, and disagreement-plus-boundary strata. Across these vectors, the within-prompt variable rate was 27.5%, the mean pairwise repeat-disagreement rate was 11.5%, and systematic disagreement vs. Exec-spec was 42.5%. The vectors partitioned into stable agreement (*n* = 54), stable systematic disagreement (*n* = 44), noise-only disagreement (*n* = 15), and mixed behavior (*n* = 7).

**Table 10:**
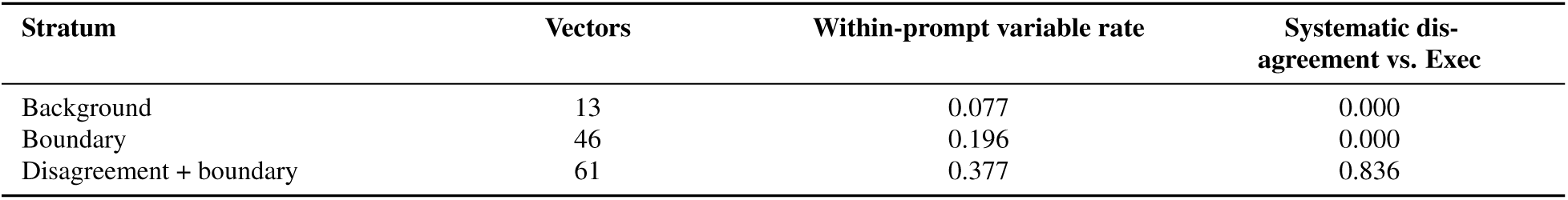
Stochastic decomposition by sampling stratum.

| Stratum | Vectors | Within-prompt variable rate | Systematic disagreement vs. Exec |
| --- | --- | --- | --- |
| Background | 13 | 0.077 | 0.000 |
| Boundary | 46 | 0.196 | 0.000 |
| Disagreement + boundary | 61 | 0.377 | 0.836 |

Stochastic variability concentrated at rule boundaries, but systematic disagreement was confined to the disagreement-boundary stratum, confirming that the bulk of variance in the full cohort is not random call-to-call noise.

